# Simultaneous T1, T2, R2*, PDFF, and Triglyceride Composition Estimation in the Head and Neck Using Magnetic Resonance Fingerprinting on a 1.5T MRI-Linear Accelerator Hybrid System: A Prospective R-IDEAL Stage 0/1 Study

**DOI:** 10.64898/2025.12.03.25341501

**Authors:** Lucas McCullum, Ergys Subashi, Samuel L. Mulder, Brian A. Taylor, Ken-Pin Hwang, Clifton D. Fuller, Jason Ostenson

**Affiliations:** UT MD Anderson Cancer Center UTHealth Houston Graduate School of Biomedical Sciences, Houston, USA; Department of Radiation Oncology, The University of Texas MD Anderson Cancer Center, Houston, TX, USA; Department of Radiation Physics, The University of Texas MD Anderson Cancer Center, Houston, TX, USA; Department of Imaging Physics, The University of Texas MD Anderson Cancer Center, Houston, TX, USA; Department of Radiology, The University of Washington, Seattle, WA

**Author notes:** Co-Corresponding Authors: Jason Ostenson < >, Clifton D. Fuller < >. **Notes:** This manuscript is the result of funding in whole or in part by the National Institutes of Health (NIH). It is subject to the NIH Public Access Policy. Through acceptance of this federal funding, NIH has been given a right to make this manuscript publicly available in PubMed Central upon the Official Date of Publication, as defined by NIH.

**Keywords:** Magnetic Resonance Fingerprinting, MR-Linac, Quantitative Imaging, Head and Neck, MRI

## Abstract

**Background and Purpose:** The extraction of quantitative biomarkers on the MR-Linac, which have been shown to be promising in the head and neck, remains a significant gap due to the strict time constraints. Therefore, the purpose of this study was to assess the technical feasibility of magnetic resonance fingerprinting (MRF) to simultaneously extract quantitative T1, T2, R2*, PDFF, and triglyceride composition in the head and neck on the MR-Linac.

**Materials and Methods:** Inversion-prepared, multi-echo, golden angle radial, variable flip angle, MRF data were acquired on a 1.5T MR-Linac. The data were corrected for phase errors in the bipolar multi-echo acquisition followed by low rank reconstruction with total variation regularization. A dictionary of signals parameterized by T1/T2 was calculated with an extended phase graph algorithm. Singular images were constructed using a signal subspace from the dictionary and the first singular images were then jointly fit for R2* and PDFF using the off-resonance information from an independent B0 map. The complete set of B0/T2*-corrected fat-water separated water singular images were fit for T1/T2. Triglyceride composition was computed using the multi-echo complex readout. Three phantoms and three healthy volunteers were assessed for accuracy, repeatability, and reproducibility against validated reference sequences across five days using a test-retest setup.

**Results:** All coefficients of determination were at least 0.9 against reference sequences in standard phantoms across all quantitative values. The repeatability and reproducibility averaged under 5% in phantoms and under 10% in healthy volunteers.

**Discussion:** The proposed MRF acquisition demonstrated the potential to replace the current time-intensive sequences on the 1.5T MR-Linac, thus increasing the adoption of specialized quantitative imaging biomarker approaches at high temporal density in the head and neck.

**Graphical Abstract:** 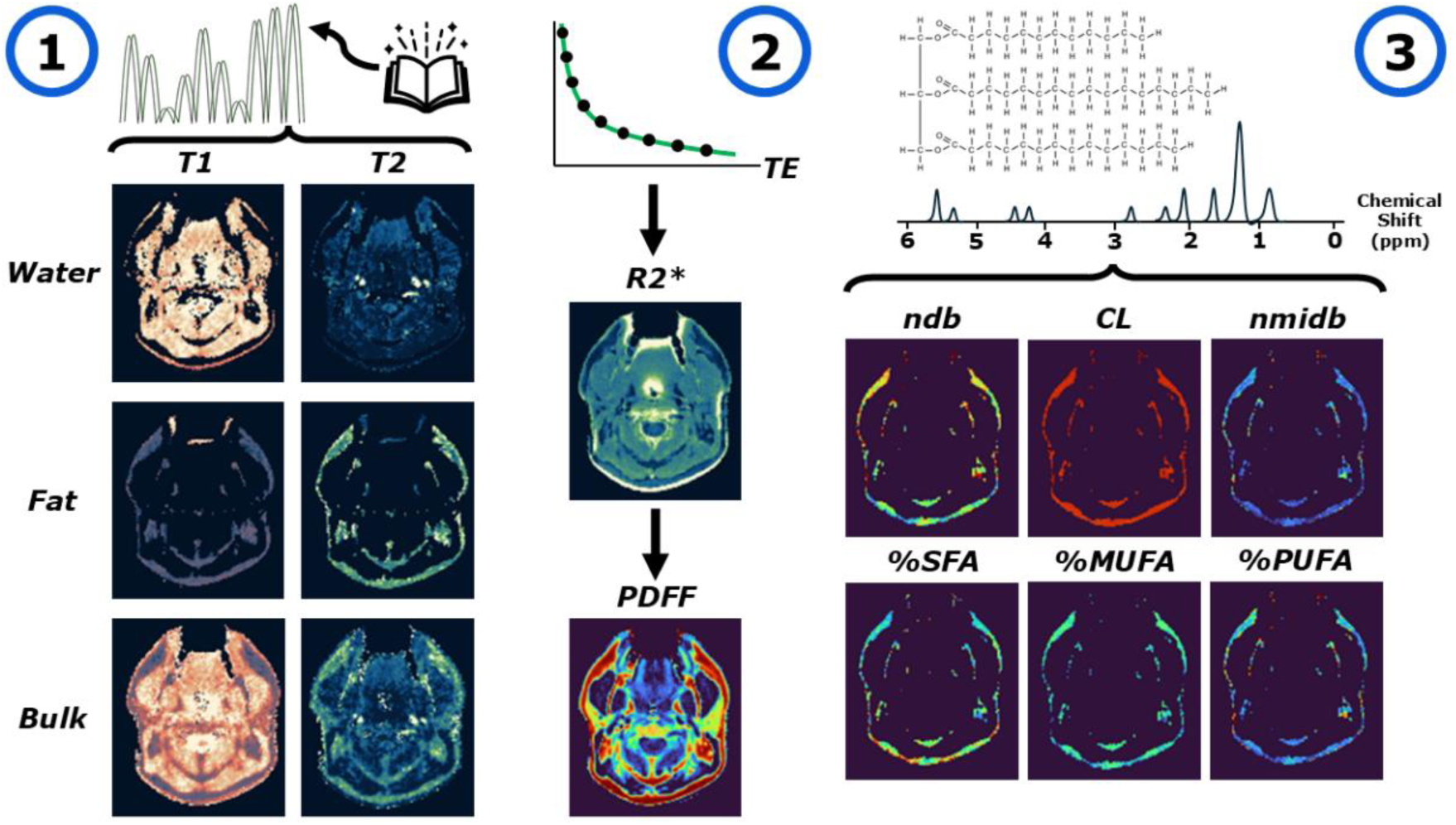

## 1. Introduction

On the MR-Linac, an integrated magnetic resonance imaging (MRI) and radiation therapy delivery linear accelerator (Linac) device^1^, head and neck cancer patients may receive daily MRI acquisitions for up to six weeks^2^ opening the door for robust normal tissue response modeling and imaging biomarker-based treatment adaptation^3^. Specifically, recent work has shown that T1-weighted signal intensity^4^ and fat fraction change in salivary glands following radiation-induced damage^5,6^. A recent study also demonstrated quantitative T1 and T2 changes in the gross tumor volume from pre- to mid- to post-radiation therapy^7^. Further work probing quantitative tissue T1, T2, apparent diffusion coefficient (ADC), and dynamic contrast enhanced (DCE) properties has recently been investigated for feasibility, demonstrating clinically significant levels of repeatability and reproducibility^8,9^. However, serial quantitative imaging suffers from prohibitively long scan times, resulting in limited sequences being available per treatment session due to time constraints for patient comfort and clinical throughput^8,10,11^. Further, acquiring quantitative maps serially will suffer from higher probability of misregistration error due to the potential for patient adjustments both intra- and inter-scan^12^. Therefore, simultaneous multiparametric quantitative MRI methods must be explored to fully take advantage of the limited time available on the MR-Linac.

Prior studies have evaluated the technical feasibility of the 2D multi-dynamic multi-echo (MDME; SyntheticMR AB; Linköping, Sweden) sequence for simultaneous quantification of T1, T2, and PD in a scan time under six minutes using the 1.5T MR-Linac^13,14^. Additionally, the multi-echo Dixon sequence (i.e., mDIXON-Quant on Philips systems) can provide quantitative B0, R2*, and PDFF maps from a single acquisition^15^. The resulting individual echo images in complex format can also be used to estimate the composition of triglycerides^16^. Magnetic resonance fingerprinting (MRF), on the other hand, is a recent innovation designed to simultaneously extract multiple quantitative property maps in a single, co-registered, acquisition^17^. Some examples^18^ are mapping T1 and T2^17^, T1_ρ_^19^, B_0_ and B_1_ ^+20^, R2*^21^, cerebral blood volume^22^, diffusion^23^, perfusion^24^, and fat signal fraction^25,26^. However, on the MR-Linac, only four studies to the author’s knowledge have evaluated the potential of MRF on the MR-Linac clinical workflow^11,27–29^ with only one focused on the 1.5T MR-Linac^11^ which investigated T1, T2 and PD.

While promising, the integration of quantitative maps currently available clinically (i.e., MDME for T1, T2, and PD; mDIXON-Quant for B_0_, R2*, PDFF, and triglyceride composition) into a single acquisition remains unexplored on the 1.5T MR-Linac. Therefore, the purpose of this study is to investigate the technical feasibility of utilizing MRF on the 1.5T MR-Linac to simultaneously quantify T1, T2, R2*, PDFF, and the derived triglyceride composition in the head and neck region as a first-in-humans R-IDEAL^30^ Stage 0/1 study. This will be achieved and shown by (1) validating quantitative accuracy, repeatability, and reproducibility in phantoms, and (2) validating quantitative accuracy, repeatability, and reproducibility in healthy volunteers. The results of this study can be used to determine the threshold for true quantitative changes outside of acquisition variability using our proposed MRF technique in head and neck cancer patients treated on the 1.5T MR-Linac.

## 2. Methods and Materials

### 2.1. MRI Acquisition Parameters

All data was acquired using a 1.5T (Philips Healthcare; Best, The Netherlands) Elekta Unity MR-Linac (Elekta AB; Stockholm, Sweden) running software version R5.7.1.2 and using the 2 x 4-channel anterior/posterior coils in place of the available quadrature body coil (QBC). For reference, this MR-Linac has a maximum gradient strength of 15 mT/m and slew rate of 65 T/m/s.

The proposed MRF sequence utilized a golden angle (∼111°) full-spoke radial gradient echo acquisition, 100 ms inversion time pre-pulse, and a nine-echo readout. A total of 2048 dynamics were acquired where the flip angle was varied between 0° and 60° by a series of squared sine waves with pseudo-random varying amplitude while the TR was kept constant at 20 ms. A first-order shim^31^ was applied to optimize the B_0_ magnetic field homogeneity while a high time bandwidth product of 10 was chosen for the RF pulse with a sinc-gauss shape to avoid slice profile B_1_^+^ heterogeneity effects using a 2D approach^32^. A total of eight crusher gradient cycles were applied between successive RF pulses to ensure full residual magnetization dephasing to prevent signal contamination^25,33,34^. Additionally, an extended waiting time was applied at the beginning of the sequence to ensure all magnetization fully recovered and is known to be in equilibrium from any previous MRI scans.

To validate the proposed MRF quantification accuracy, we developed independent quantitative mapping sequences and validated their performance in phantoms with known reference values. For quantitative T1 and T2, the 2D multi-dynamic multi-echo (MDME; SyntheticMR AB; Linköping, Sweden) sequence was used as previously rigorously validated on the 1.5T MR-Linac^13,14^. The quantitative R2* and PDFF values were validated using the mDIXON-Quant^6,35^ technique which employs a multi-echo gradient echo sequence and multi-peak fat model with eddy current compensation post-processing in real-time on the scanner. Immediate calculated R2* and PDFF maps were available from the scanner in DICOM format^36^. A dual-echo independent B_0_ map was acquired for off resonance corrections. The total scan time for all acquisitions was 4 minutes 22 seconds per repetition. Further acquisition details for both the proposed MRF acquisition and reference acquisitions can be seen in **Table 1**.

**Table 1.** The MRI acquisition parameters for the proposed MRF acquisition and its related validation sequences. Note that some parameters may not be valid for the specified sequence which is indicated by a dashed line. Abbreviations: N_dynamics_ = number of dynamics, NSA = number of signal averages, TE = echo time, TI = inversion time, TR = repetition time, TSE = turbo spin echo.

|  | **B<sub>0</sub> Map** | **mDIXON-Quant** | **MDME** | **MRF** |
| --- | --- | --- | --- | --- |
| Field of View (mm<sup>3</sup>) | 256 x 256 x 10 | 256 x 256 x 30 | 256 x 256 x 45 | 256 x 256 x 10 |
| Acquired Voxel Size (mm<sup>3</sup>) | 1.5 x 1.5 x 10 | 2.4 x 2.4 x 10 | 2 x 2 x 3 | 1.5 x 1.5 x 10 |
| Matrix Size | 172 x 172 x 1 | 108 x 107 x 3 | 128 x 117 x 15 | 172 x 172 x 1 |
| Reconstructed Voxel Size (mm<sup>3</sup>) | 0.8 x 0.8 x 10 | 0.8 x 0.8 x 10 | 0.8 x 0.8 x 3 | 0.8 x 0.8 x 10 |
| Flip Angle (°) | 20 | 5 | 160 | 0 – 60 (variable) |
| TR (ms) | 12 | 20 | 4000 | 20 |
| TE (ms) | 4.6 | 1.49 | 24 / 127 | 3.21 |
| ΔTE (ms) | 4.6 | 1.19 | - | 1.53 |
| $N_{\text{TE}}$ | 2 | 6 | 2 | 9 |
| Bandwidth (Hz/Pixel) | 734.1 / 0.296 | 1512.9 / 0.144 | 230.9 / 0.941 | 949.9 / 0.229 |
| NSA | 5 | 4 | 1 | 1 |
| Additional Notes | - | - | CS-SENSE = 2,<br>TSE Factor = 12 | $N_{\text{dynamics}}$ = 2048,<br>TI = 100 ms |
| Scan Time (m:ss) | 0:21 | 0:24 | 2:44 | 0:53 |

### 2.2. Phantom / Human Subject Assessment

The CaliberMRI “ISMRM/NIST” Premium System Phantom Model 130 phantom (CaliberMRI; Boulder, CO, USA) was used as a reference for NIST-traceable T1 and T2 values^37^. The T2 vial layer was utilized to validate quantitative T1 and T2 parameter concordance due to their more clinically relevant T2 values. Another phantom was developed in-house for PDFF concordance using varying percentages of distilled water, vegetable oil (100% soybean oil), and a liquid lecithin emulsifier^38,39^. Nominal fat fractions of 0%, 25%, 50%, 75%, and 100% were created in 30 mm diameter / 50 mL vials (SimPure; Auburn, WA, USA) and partially submerged in distilled water for better signal characterization due to their smaller size. The vials were shaken immediately prior to scanning to ensure full fat / water mixing. Since the R2* did not have reported nominal values for the ISMRM/NIST phantom or our in-house phantom, we additionally scanned the Calimetrix Model 725 PDFF-R2* phantom^40^ (Calimetrix; Madison, WI, USA).

The bore temperature was monitored using a manual thermometer which regularly read near 20°C across all acquisitions for each scanner. Repeatability and reproducibility in the ISMRM/NIST phantom, Calimetrix phantom, and the in-house PDFF phantom were conducted by scanning them for a total of five days in a row using a test-retest methodology for each session. Specifically, the phantom was re-positioned following each scan, thus necessitating novel parameters for frequency adjustment, B_0_ shimming, B_1_^+^ calibration, and coil tuning. This was done separately for each phantom. Quantitative analysis was restricted to the following clinical ranges^41^: T1 = 250 – 1750 ms, T2 = 25 – 200 ms, PDFF = 0 – 100%, and R2* = 0 – 100 1/s.

In addition to phantom evaluation, a total of three healthy subjects were recruited and scanned using the imaging protocol in **Table 1** in a test-retest style where they were asked to leave the bore and move around prior to re-positioning to trigger the new pre-scan parameters. All participants provided written informed consent to an internal volunteer imaging protocol (PA15-0418), both approved by the institutional review board at The University of Texas MD Anderson Cancer Center. The desired structures of interest in the head and neck that were assessed were the bilateral parotid glands (PG), bilateral masseter muscle (MM), bilateral medial pterygoid muscle (MPM), and bilateral middle superficial cheek fat (MSCF). Each structure was contoured for each timepoint and repetition using the individual gradient echo images from the B_0_ map by an experienced clinical professional in 3D Slicer^42^ (https://www.slicer.org).

### 2.3. MRF Dictionary Generation and Image Reconstruction

The MRF reconstruction was partially completed by a pre-computed dictionary which was generated using MATLAB R2023a (The MathWorks, Inc.; Natick, MA, USA). The flip angle train was read using comma separated value (CSV) format and simulated using an extended phase graph algorithm^43^. The simulated T1 values ranged from 100 – 2000 ms with 75 log-spaced steps and the simulated T2 values ranged from 5 – 500 ms with 75 log-spaced steps. The total computational time for dictionary generation was 42 sec using a Dell Inc. Latitude 7320 operating Windows 10 Enterprise with an 11^th^ Gen Intel Core i7-1185G7 processor with 4 cores and 16 GB of RAM. The wide range of T2 values was to accommodate the ISMRM/NIST phantom which includes large T2 values along with its large T1 value vials.

The first step in the MRF reconstruction was the processing of the raw image data which was saved directly from the scanner using Philips DATA/LIST format. Next, the inter-excitation basis for the T1/T2 dictionary was calculated from singular value decomposition^44^ using a rank factor of 0.9999 (i.e., components contributing to the highest 99.99% of the total energy were kept). These reduced number of components were used to generate a compressed dictionary for later signal matching. Each echo, and corresponding singular images, was reconstructed using the forward and adjoint Flatiron Institute nonuniform fast Fourier transform^45,46^ (NUFFT) operators at a precision of 10^-10^ with low-rank subspace dimensionality reduction^47^ from the previously determined number of significant components via a low-rank total variation^48^ (LRTV) reconstruction using 40 iterations, initial step size = 2^-10^, regularization weight = 5 x 10^-3^, and backtracking adaptive step size search. The coil sensitivity maps were then computed from the LRTV reconstruction using a matched filtering^49^ approach from the first singular image of the first echo and then applied to subsequent echoes to correct for inhomogeneous signal intensities.

To achieve estimated R2* and PDFF, the reconstructed first singular images of all echoes and the independent B_0_ images were processed through the PRESCO algorithm^50^ assuming 2.8 double bonds, non-negative R2* and PDFF, a maximum of 10 iterations for fat-water swap reduction, 8 iterations for the gradient descent, and 3 iterations for the line search. To estimate T1 and T2, the separated fat and water first singular image components were individually fit to the pre-computed and compressed dictionary using matched filtering^51^. Since our T1 and T2 estimates were separated into fat and water components, we utilized the bulk estimation (weighted sum of fat- and water-separated first singular images according to the fat signal fraction) to compare to our ground truth references since they were not fat or water suppressed prior to image acquisition.

Then, to estimate the complex fat-water separated signal components, the reconstructed singular images of all echoes was first corrected for using the independent B_0_ and phase maps jointly with the R2* map from PRESCO and then separated using pre-defined 11-peak fat and water signal models^52^ and recombined using a weighted sum of all singular images. Then, to compute the number of double bonds (ndb) and number of methylene interrupted double bonds (nmidb), the ratio between the signal from both the olefinic and diallylic peaks and the bulk methylene peak were determined using the signal evolution of the first singular image and a system of equations of the theoretical peak amplitudes to solve for ndb and nmidb from Peterson et al. 2021^53^. Next, the chain length (CL) was computed from Bydder et al.^16^ and the fraction of saturated fatty acids (%SFA), monounsaturated fatty acids (%MUFA), and polyunsaturated fatty acids (%PUFA) from Peterson et al.^53^. The fraction of triunsaturated fatty acids (%TUFA) was assumed to be 0% and ignored due to its relatively low potential bias at 2% fixed across the entire volume^54,55^. Soybean oil reference values for ndb and nmidb were derived from Peterson et al. 2013^56^ and the %SFA, %MUFA, and %PUFA were derived from the United States Department of Agriculture Nutrient Database. An abbreviated visual methods overview of the methods is provided in **Figure 1**.

**Figure 1.**
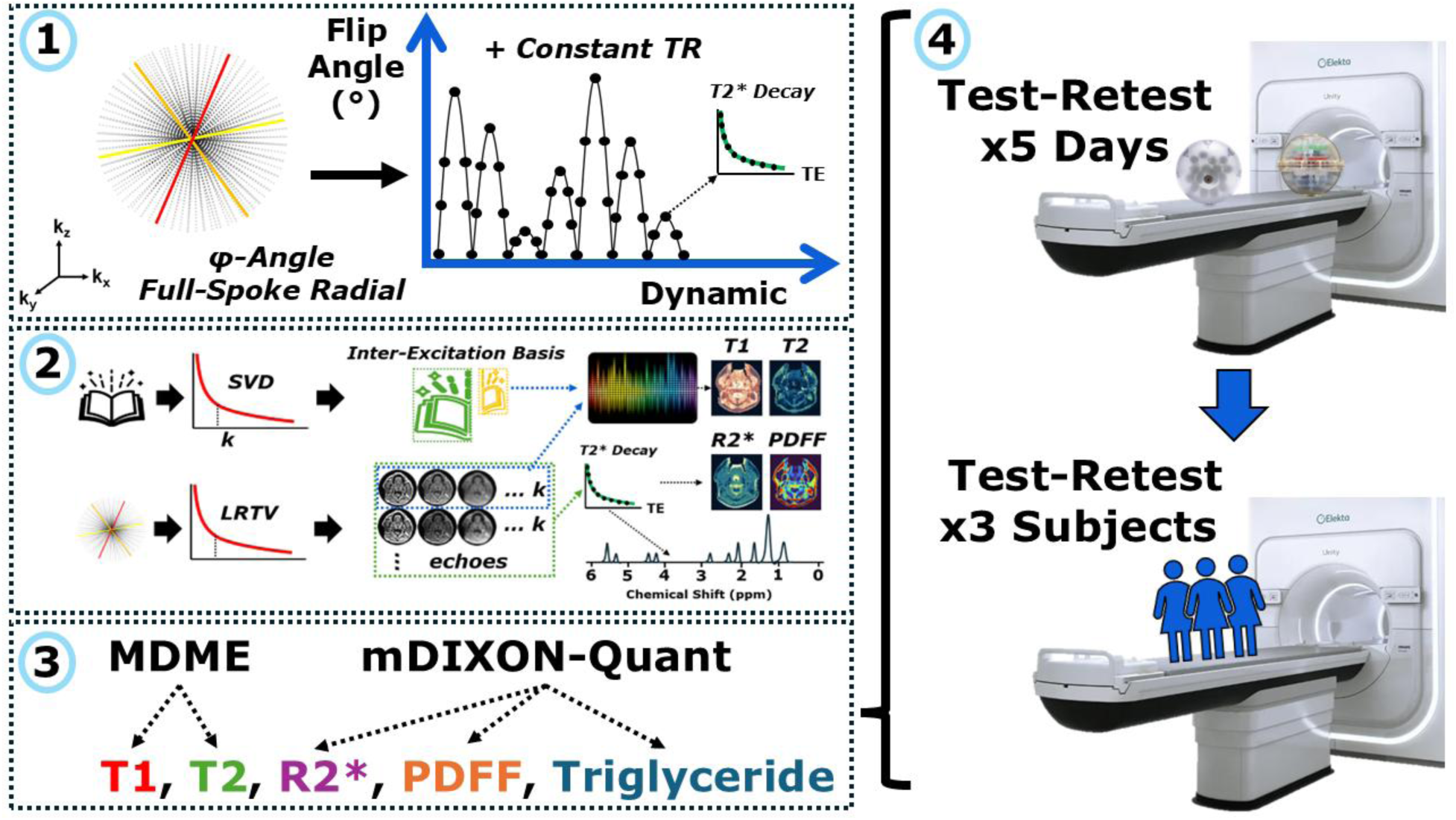
A high-level graphical overview of the MRF methodology used in this study: 1) image acquisition using a golden angle radial readout across multiple dynamics with variable flip angle and constant TR, 2) image reconstruction using a LRTV-based reconstruction followed by dictionary matching for T1/T2 and curve fitting for R2*/PDFF/triglyceride composition, 3) comparison to validated references, and 4) test-retest studies in phantoms and human subjects.

### 2.4. Statistical Analysis

All relevant analysis concerning statistical methods were formulated using the guidelines for reporting Statistical Analyses and Methods in the Published Literature (SAMPL)^57^. First, the concordance between the reference sequences and the phantom nominal values were compared using the squared Pearson correlation coefficient^58^, or the coefficient of determination (r^2^), for the T1, T2, R2*, and PDFF values. Next, a linear fit for the reference acquisitions against nominal phantom reference values was computed from solving a Vandermonde matrix^59^. For this computation, a y-intercept of 0 was chosen to assume that a quantitative reference value of 0 will lead to a direct matching to the proposed MRF values due to no detectable signal (T1, T2, R2*) and no confounding effects of fat (PDFF). The resulting first-order slopes were extracted and used to determine linear bias. After establishing the linear bias in the reference sequences, the proposed MRF sequence values against the reference sequence values were compared using the same strategy along with a Bland-Altman^60^ analysis. To validate the ndb, nmidb, CL, %SFA, %MUFA, and %PUFA, the 100% soybean oil vial was compared between all the repeated acquisitions and the known reference values^56,61^ and the United States Department of Agriculture Nutrient Database. The median and interquartile range (IQR) was used to assess its agreement to these reference values. The repeatability and reproducibility of both the proposed MRF sequence and the reference sequences were computed using the percentage of the standard deviation to the mean, or the coefficient of variation (CoV).

## 3. Results

The performance of MDME for T1 and T2 and mDIXON-Quant for R2* and PDFF across the five repeated days of test-retest in their respective reference phantoms are shown in **Figure 2**. The MDME sequence showed an r^2^ agreement of 0.99 for T1 and 0.99 for T2 while the slope for each linear fit was 1.08 and 0.87, respectively. Similar results were seen using the mDIXON-Quant sequence with an r^2^ agreement of 0.94 for R2* and 0.99 for PDFF with slopes of 1.02 and 1.00, respectively.

**Figure 2.**
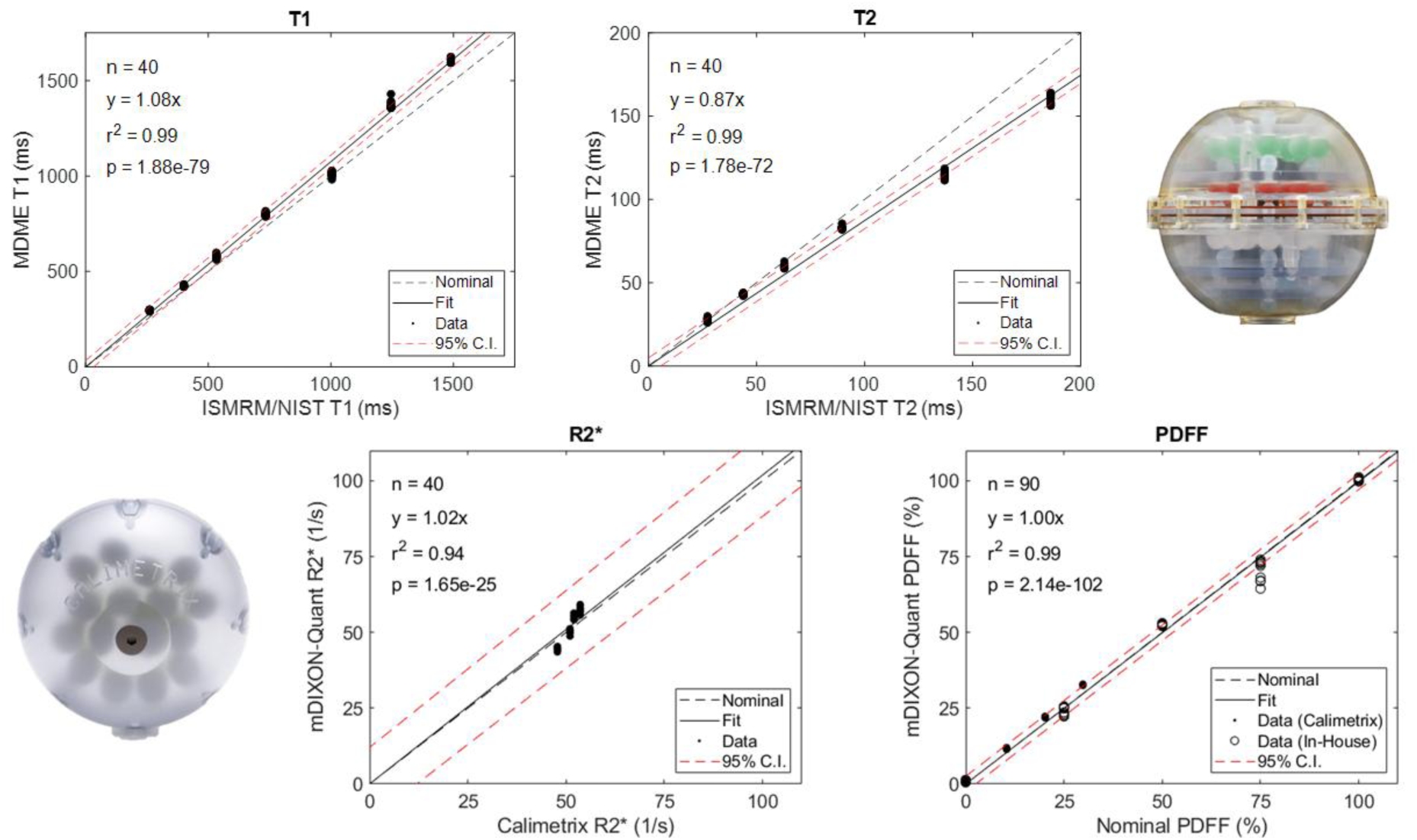
Concordance plots of the quantitative T1 (top left), T2 (top right), R2* (bottom left), and PDFF (bottom right) values from the proposed ground-truth sequences (MDME for T1 and T2, mDIXON-Quant for R2* and PDFF) compared to the phantom nominal reference values (dashed black line, CaliberMRI ISMRM/NIST Model 130 for T1 and T2, Calimetrix Model 725 for PDFF and R2*, and in-house phantom for PDFF). For each series of data, a linear line of best fit (solid black line) was computed along with its 95% confidence intervals (dashed red lines).

After verification of the reference sequences, direct comparison between these sequences and the MRF values in the phantoms could be made, allowing for future comparison in human subjects. The results comparing MDME (T1 and T2) and mDIXON-Quant (R2* and PDFF) with MRF values across the five repeated days of test-retest are shown in **Figure 3**. The r^2^ agreements were 0.99, 0.94, 0.93, and 0.98 while the linear fit slopes were 0.98, 1.14, 1.28, and 1.08 for T1, T2, R2*, and PDFF, respectively. A direct comparison and Bland-Altman analysis of the proposed MRF values against the phantom references values is shown in **Figure S1** and **Figure S2**, respectively, in the **Supplementary Materials**.

**Figure 3.**
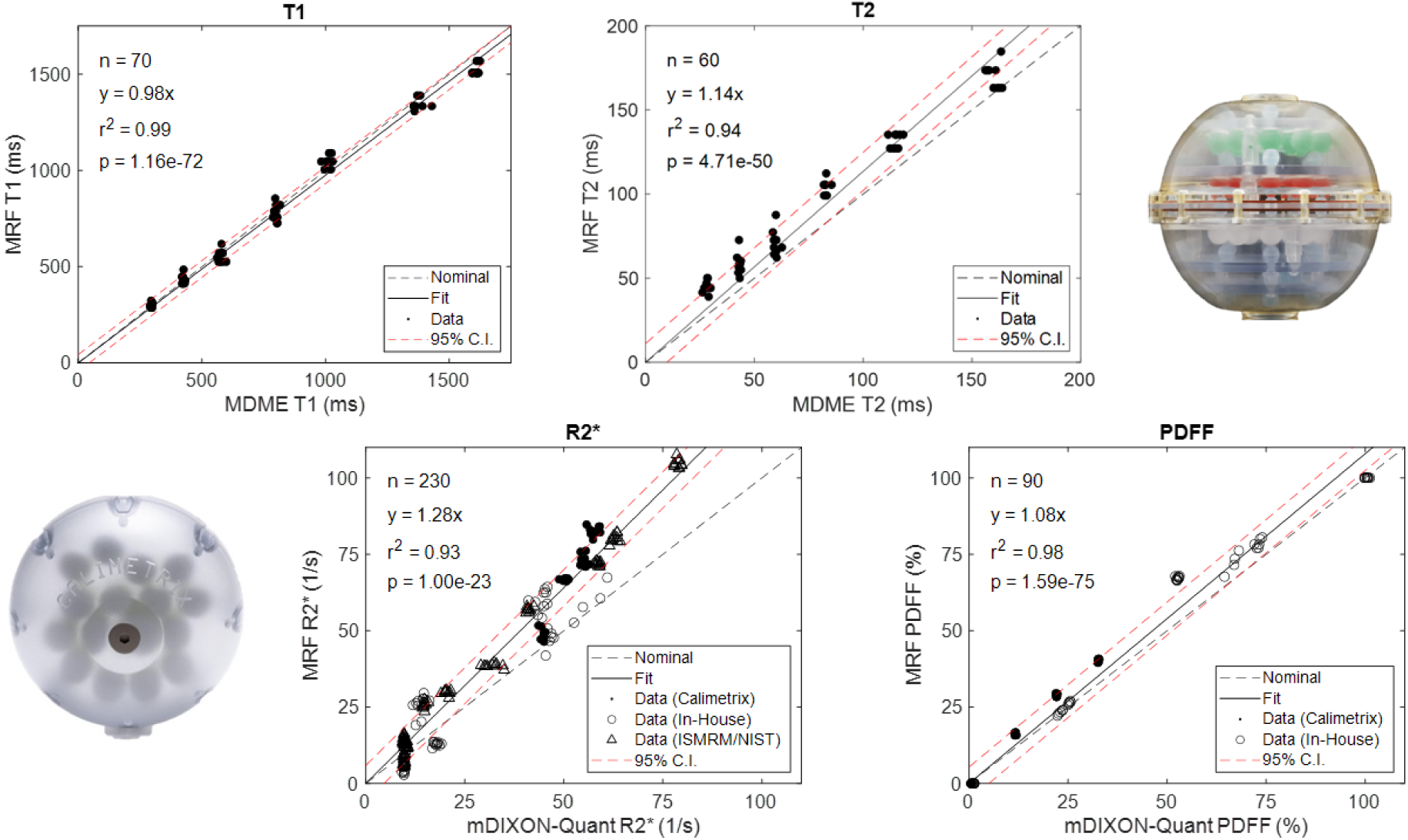
Concordance plots of the quantitative T1 (top left), T2 (top right), R2* (bottom left), and PDFF (bottom right) values from the proposed MRF sequence (black dots) compared to the reference sequences (dashed black line). For each series of data, a linear line of best fit (solid black line) was computed along with its 95% confidence intervals (dashed red lines).

For conciseness, the triglyceride composition results for both the phantom and healthy volunteers are combined in **Figure 4**. In the phantom, the ndb and nmidb were slightly underestimated while the remaining metrics were similar to soybean oil reference values. Quantitatively, the median [IQR] was 4.34 [3.98 – 4.47], 1.79 [1.53 – 1.89], 18.02 [17.91 – 18.05], 15.07 [14.32 – 18.37], 25.85 [23.16 – 31.22], and 59.74 [51.10 – 63.07] compared to reference values of 4.59, 2.07, 17.80, 15.75, 23.36, and 60.89 for ndb, nmidb, CL, %SFA, %MUFA, and %PUFA, respectively.

**Figure 4.**
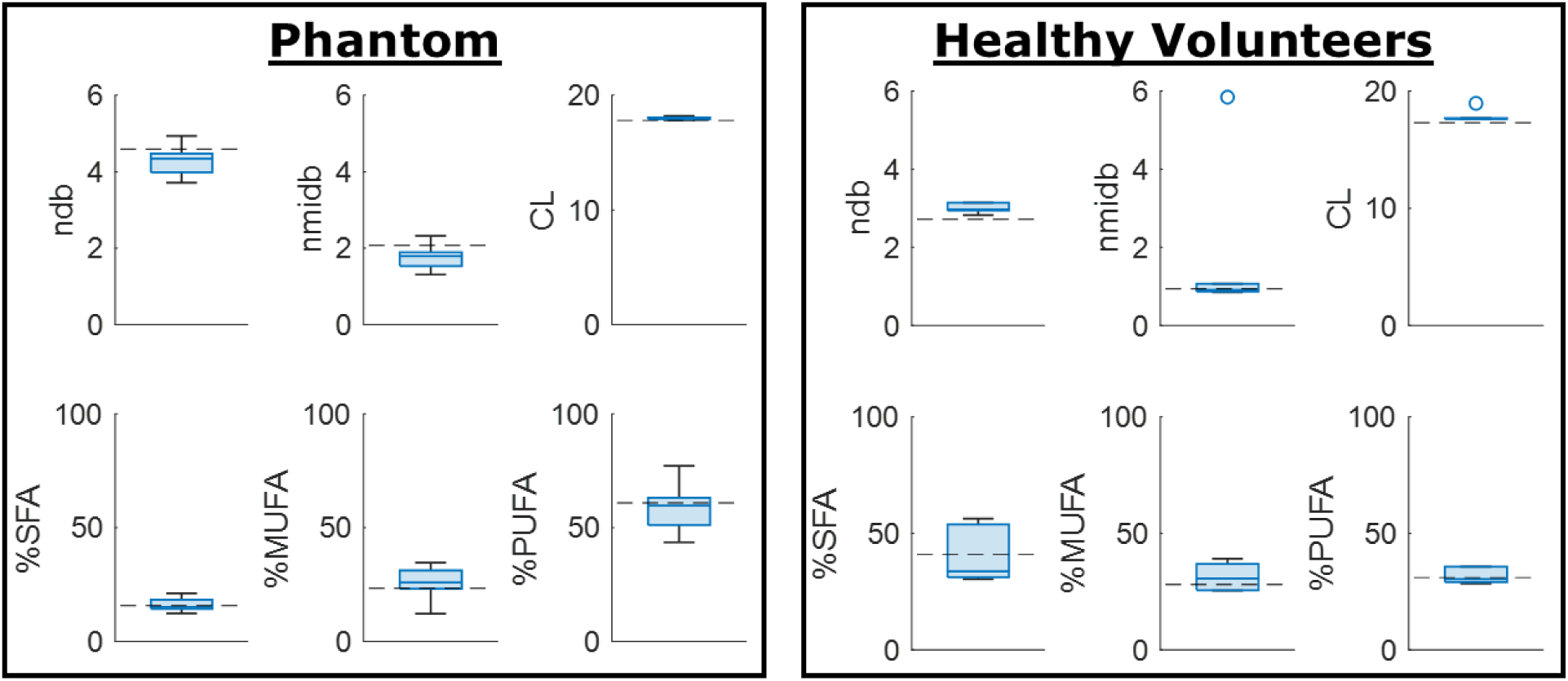
Repeatability of the triglyceride characterization in the 100% soybean oil vial from the in-house phantom (left) and the middle superficial cheek fat (right) compared to reference values (dashed black line). Note, the difference in the reference values between soybean oil and human subcutaneous adipose tissue.

In the middle superficial cheek fat in healthy volunteers, the ndb and CL were slightly overestimated while the remaining metrics were similar to reference values for subcutaneous adipose tissue. Quantitatively, the median [IQR] was 2.97 [2.95 – 3.15], 0.91 [0.87 – 1.07], 17.63 [17.62 – 17.68], 33.63 [31.11 – 53.72], 30.55 [25.53 – 36.83], and 30.37 [28.99 – 35.62] compared to reference values of 2.72, 0.94, 17.30, 41.00, 28.00, and 31.00 for ndb, nmidb, CL, %SFA, %MUFA, and %PUFA, respectively.

Since MRF acquisitions were performed across five repeated days in a test-retest format for each day, the CoV could be characterized for both repeatability and reproducibility and is shown in **Figure 5**. All CoV were within 10% except for the reproducibility of the lowest R2* values which were ∼11%. The lowest CoV was seen in the T1 values >1000 ms, T2 values >75 ms, R2* values >20 1/s except for ∼50 1/s, and PDFF values of 50% and 100%.

**Figure 5.**
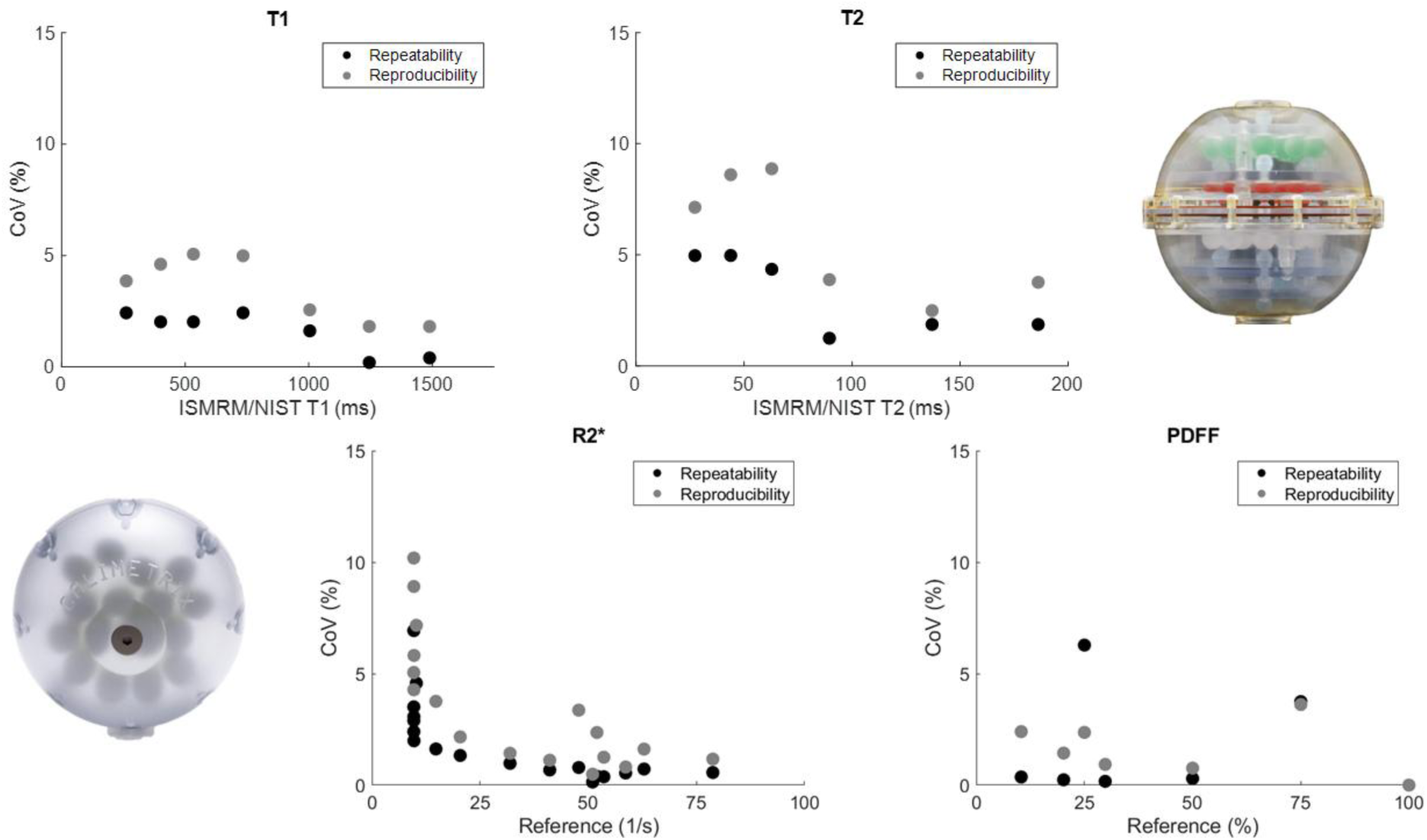
Repeatability and reproducibility plots of the quantitative T1 (top left), T2 (top right), R2* (bottom left), and PDFF (bottom right) values from the proposed MRF sequence compared to the nominal values (ISMRM/NIST phantom reference values for T1/T2 and Calimetrix phantom for R2*/PDFF) or reference sequences (mDIXON-Quant for R2*). Note, the repeatability and reproducibility of 0% for PDFF levels of 0% and 100% due to it all being the same value.

A representative compilation of the quantitative maps in a healthy volunteer generated from the proposed MRF sequence including T1 (fat / water / bulk), T2 (fat / water / bulk), R2*, PDFF, ndb, nmidb, CL, %SFA, %MUFA, and %PUFA is shown in **Figure 6**.

**Figure 6.**
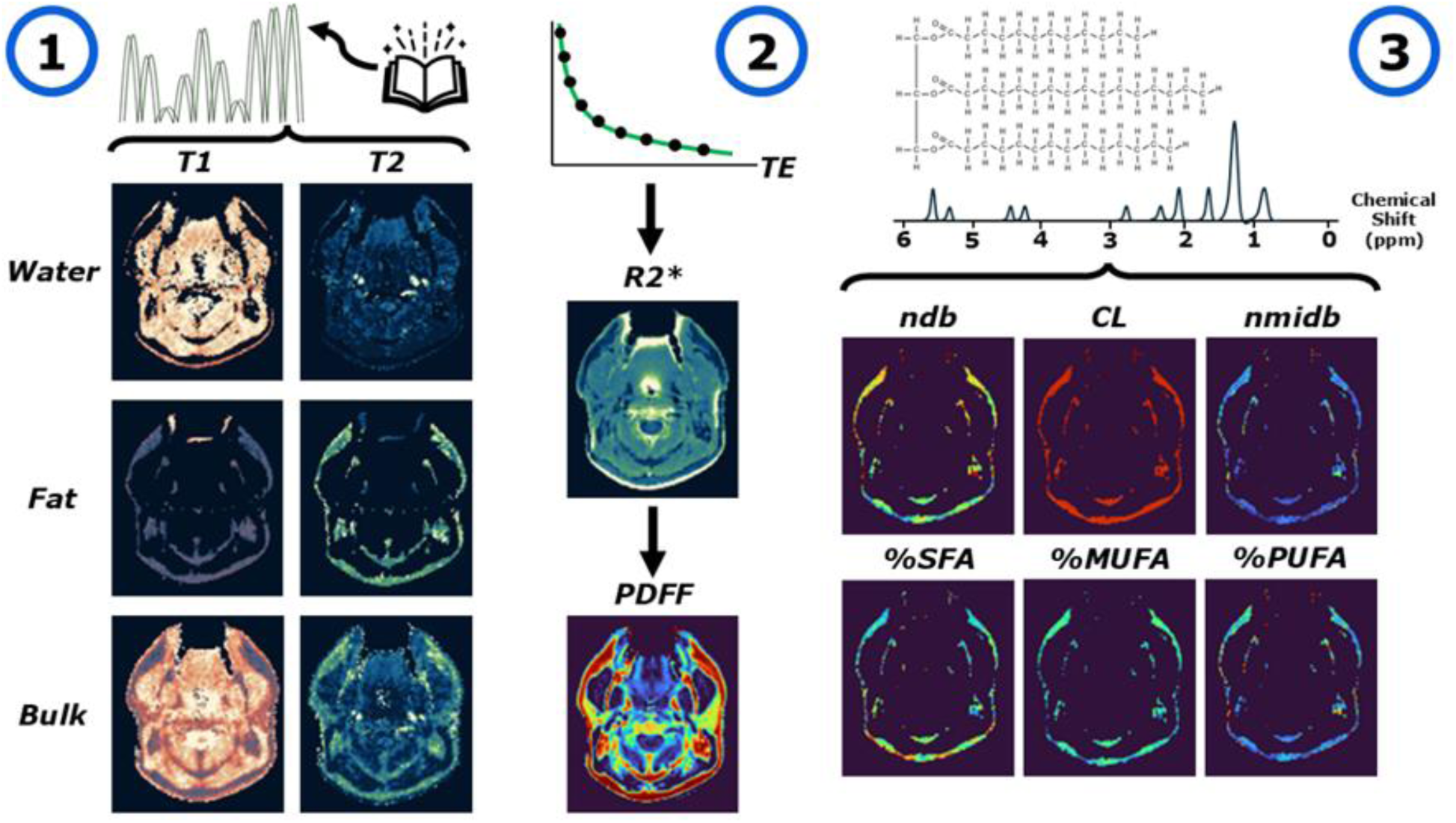
A visual summary of all the quantitative maps acquired in a single acquisition using the proposed MRF sequence in a healthy volunteer with a permanent retainer leaving an anterior signal void. (1) Dictionary matching is utilized to fit for T1 and T2 which can then be separated into both fat and water components. (2) Curve fitting of the gradient echo signal decay can extract both the decay rate, R2*, and percent fat fraction, PDFF. (3) Further fitting of the complex gradient echo signal decay to various fat spectral peaks can lead to extraction of the ndb, nmidb, CL, %SFA, %MUFA, and %PUFA. Note, a fat mask (MRF PDFF > 90%) is used for the triglyceride characterization maps due to their unreliability in regions with minimal fat content.

A comparison of the quantitative values in each of the contoured structures across both test-retest scans in the three healthy volunteers for the proposed MRF acquisition against the reference acquisitions is shown in **Figure 7**. The T1 values showed the best agreement with an r^2^ of 0.84 and slope of 1.05 while the T2 values were slightly overestimated, though consistent, with an r^2^ of 0.76 and slope of 1.16 compared to the reference MDME acquisition. The R2* values showed the worst agreement with an r^2^ of 0.47 and slope of 0.80, likely due to the drop in accuracy at the higher R2* values (>50 1/s). Finally, the PDFF values were the most consistent with an r^2^ of 0.98 and slope of 1.22.

**Figure 7.**
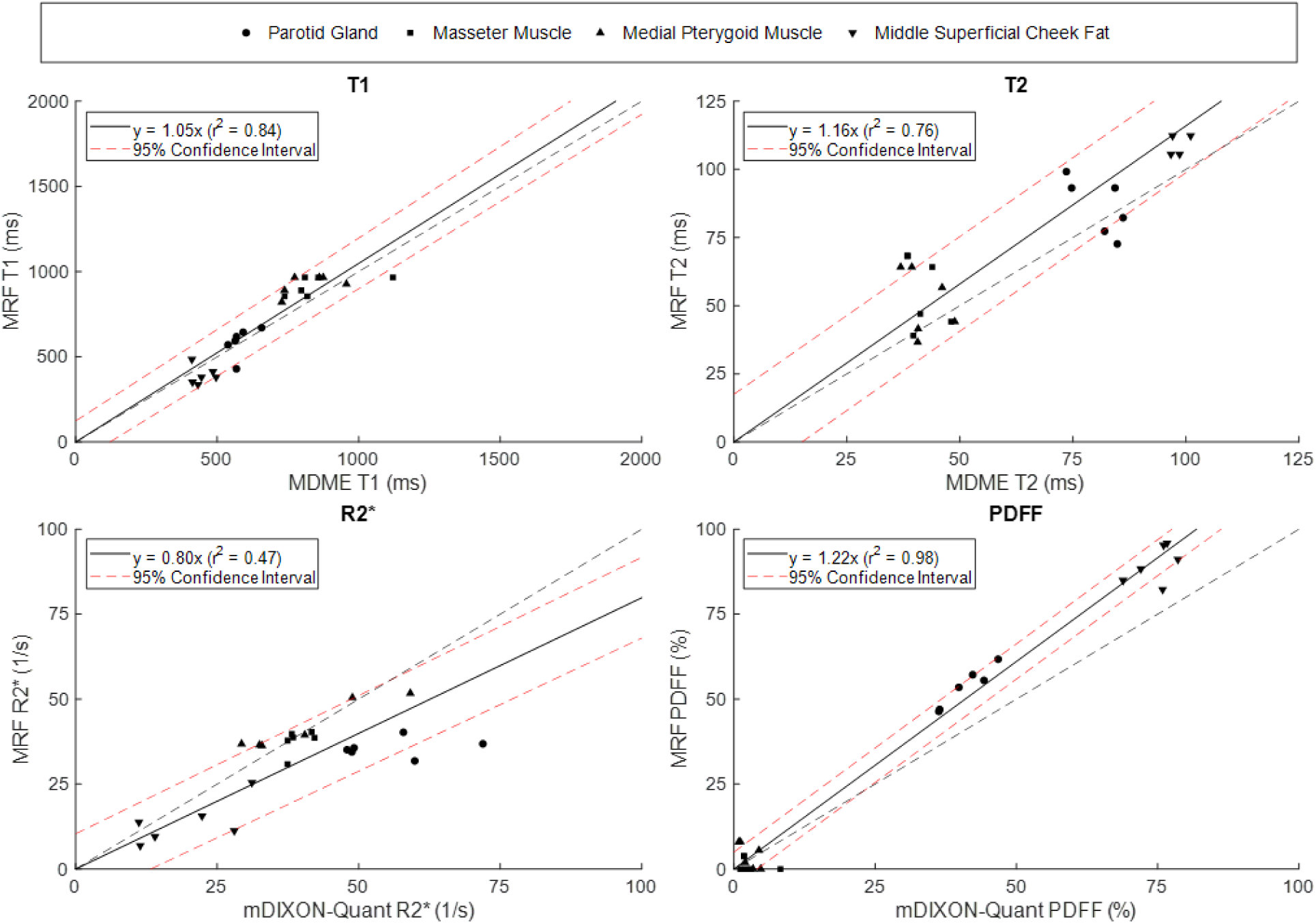
A comparison between the proposed MRF acquisition and the reference acquisition (MDME for T1 / T2 and mDIXON-Quant for R2* / PDFF) across the two test-retest scans in the three healthy volunteers.

The quantitative repeatability and reproducibility results for **Figure 7** are shown in **Figure 8** with the addition of the triglyceride composition analysis. A lower CoV was seen in the MRF acquisition compared to MDME for both muscles in the quantitative T1 values. However, the CoV for the MRF acquisition was higher than MDME in the quantitative T2 measurements. In most of the R2* measurements, the CoV in the MRF acquisition was lower compared to mDIXON-Quant, while being comparable for the PDFF measurements. The CoV for the repeatability and reproducibility was the lowest in the CL while higher in the nmidb with most values ranging between 10% and 30%.

**Figure 8.**
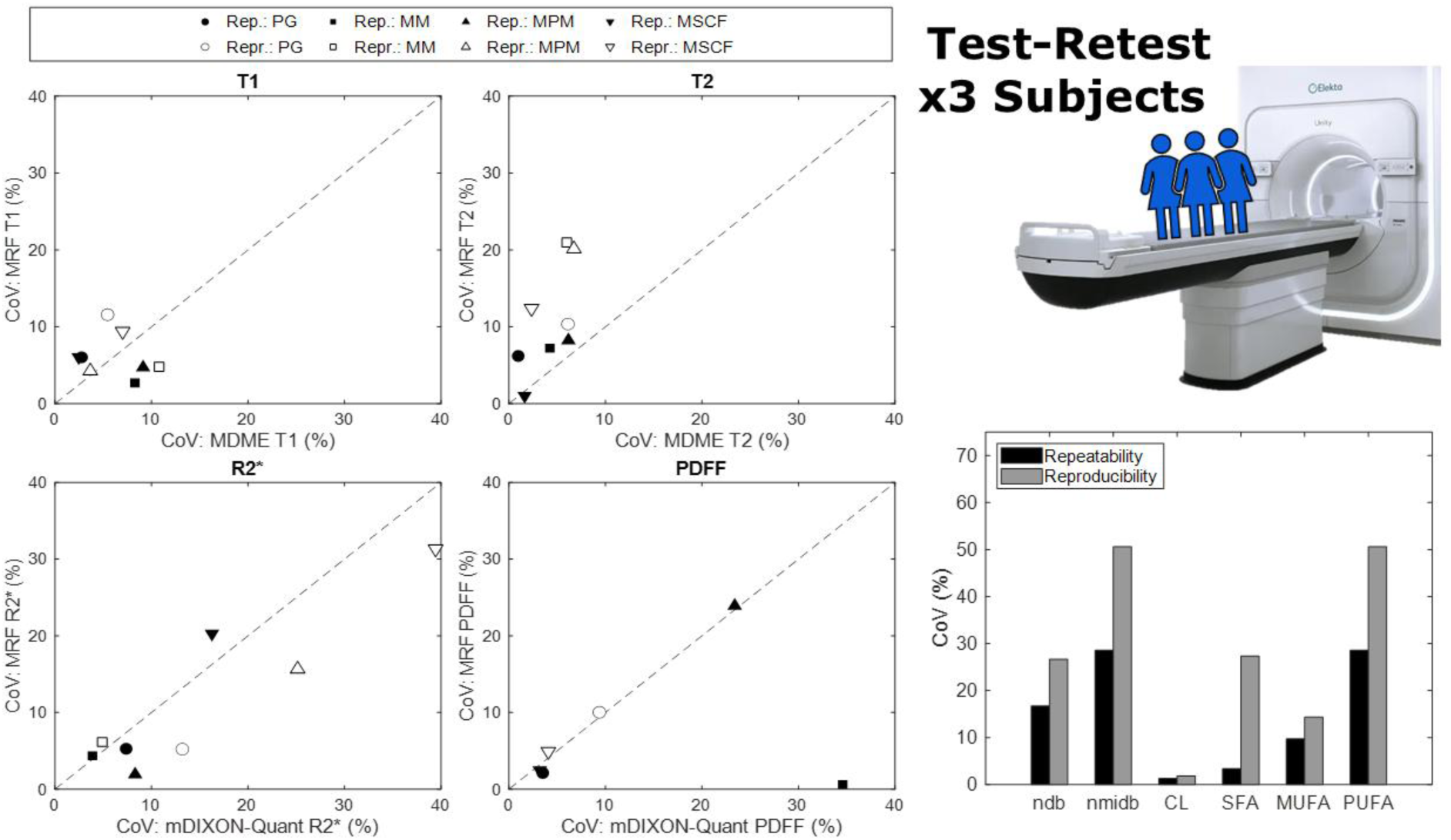
The repeatability in human subjects across the proposed MRF sequence and reference sequence for comparison for T1 (top left), T2 (top middle), R2* (bottom left), and PDFF (bottom middle) across all contoured structures. Also shown is the repeatability and reproducibility for ndb, nmidb, CL, %SFA, %MUFA, and %PUFA for the proposed MRF sequence (bottom right) for just the middle superficial cheek fat due to its high fat content. Abbreviations: rep = repeatability, repr = reproducibility, PG = parotid gland, MM = masseter muscle, MPM = medial pterygoid muscle, MSCF = middle superficial cheek fat.

## 4. Discussion

### 4.1. Summary

This work investigated the technical feasibility of simultaneously quantifying T1, T2, R2*, PDFF, triglyceride composition estimation (ndb, nmidb, CL, %SFA, %MUFA, and %PUFA) using magnetic resonance fingerprinting on a 1.5T MR-Linac. This study includes several novel contributions to the field of image-guided radiation therapy on a 1.5T MR-Linac including: (1) the first published implementation of MRF on the 1.5T MR-Linac including R2*, PDFF, and triglyceride composition, (2) the first published implementation of MRF to include structures in the head and neck, and (3) the first published evaluation of the performance of mDIXON-Quant on the 1.5T MR-Linac.

We found that our proposed MRF sequence generated quantitative T1^13,62^ and PDFF^63–65^ values agreed with published reference values while R2*^66^ was underestimated in the parotid glands whereas, in the muscles, quantitative T1^13^, T2^13^, and PDFF^65^ were in agreement with published reference values. Similar to the parotid, in the fat tissue, the quantitative T1^13^ and PDFF^64,65^ agreed with published reference values while T2^13^ was slightly overestimated. Exact quantitative R2* measurements in the muscles and fat of the head and neck R2* could not be found in existing literature. An interesting finding was that the mDIXON-Quant appeared to underestimate the PDFF in the middle superficial cheek fat which needs to be investigated further. For the triglyceride composition estimation, the IQR was within the expected reference values in the phantom except for the ndb and nmidb which were slightly underestimated and CL which was slightly overestimated. In the healthy volunteers, the ndb and CL were slightly overestimated.

### 4.2. Limitations

Several acquisition and reconstruction factors can potentially limit the precision of the quantitative measurements from MRF including patient motion, low SNR due to scan time constraints, or non-robust reconstruction pipelines. On the acquisition side, radial off-resonance effects cause phase inconsistencies across the radial spokes, especially near the end of the spoke with high frequency information, which can cause blurring, streaks, or rings^67,68^, however, our MRF acquisition utilized a high bandwidth per pixel which should minimize these concerns with the tradeoff of lower SNR. Since we limited our MRF dictionary to a maximum T1 and T2 of 2000 ms, this can potentially lead to an underestimation of free-water signal dominant materials such as the cerebrospinal fluid (CSF)^69^. Similarly, tissues with short T2 times such as bone and tendons may experience a similar effect but as an overestimation^69^. Our dictionary range and spacing was chosen with computational memory constraints in mind so more investigation should be done by increasing the range and number of intervals with larger amounts of computational memory. However, the current study still covers a vast majority of the tissues represented in the head and neck region.

From the quantitative parameter validation perspective, there are several steps which can be taken to ensure more robustness. First, the Calimetrix phantom used in this study only contained R2* values near 50 1/s in our selected clinical range of 0 – 100 1/s which limits the validation of mDIXON-Quant compared to nominal values. The dedicated 12-vial R2* phantom from Calimetrix includes values of 15, 25, 50, 75, and 100 1/s which would provide a better characterization of our clinical range and should, therefore, be investigated further. Second, we noticed an overestimation of the proposed MRF sequence’s PDFF compared to that from mDIXON-Quant at a fat fraction of 50%. This could possibly be from the maximal interference from the water and fat signals, particularly occurring in cases of large phase errors^70^ or biased T2* corrections^71^. This could potentially be corrected for in future studies by applying advanced reconstruction techniques such as dual T2* correction^71^ or an independent T2* mapping technique outside of the one computed with mDIXON-Quant. Third, due to our low sensitivity for unconstrained estimates of nmidb, we decided to utilize the formulation from Peterson et al.^53^, resulting in derived values for CL, %SFA, %MUFA, and %PUFA that were dependent on the accuracy of the ndb measurement.

### 4.3. Future Work

Looking forward to the future, for quantitative T1 and T2, an interleaved spin echo and inversion recovery sequence, or MIX^72^ acquisition on Philips devices, may yield a more accurate estimation at the price of a longer scan time compared to MDME and should be investigated further. Although not investigated here, the use of fat or water spectrally selective inversion pulses^73–75^ could have been employed for the quantitative T1 and T2 mapping techniques to validate the water and fat separated quantitative T1 and T2 values. For simultaneous R2* and PDFF estimation, the PRESCO algorithm was chosen, however further reconstruction methods should be investigated to validate its measured biases compared to mDIXON-Quant beyond the differences in first echo time and echo spacing^76^. Additionally, the use of an independent B_0_ map could be removed and replaced with the one estimated by PRESCO given it has acceptable level of bias. Another area of exploration is the overestimation in the R * values from the proposed MRF method against mDIXON-Quant in phantoms, which could be due to the relatively large first echo time of 3.21 ms and larger echo spacing of 1.53 ms which fails to accurately capture the signal decay with sufficient SNR^77^. However, the overestimation is consistent with a high r^2^ and mean repeatability and reproducibility under 5% across all vials in the clinical reference range. After further investigation, the first echo time can be reduced to a minimum of 1.8 ms on the Elekta Unity MR-Linac if a hard block pulse shape is used, however, for 2D imaging, a higher time-bandwidth product pulse is desired to prevent signal cross talk and slice-profile effects that bias relaxometry estimates.

In some cases, the CoV was large especially when the mean approached 0 such as the PDFF in muscles. Larger healthy volunteer cohorts should be studied to determine if this will reduce the CoV, especially for the reproducibility quantification. Future work can also incorporate the fitted quantitative T2 values from MRF along with the derived T2* values from PRESCO to study the T2’ values and their potential as a longitudinal biomarker^78^. A random sample consensus (RANSAC) model^79^ could be used to generate a better lines-of-best-fit which are more robust to outliers and non-linear trends, especially in the R2* fit in health volunteers which noted an increasingly negative bias at higher values. Individuals with lower amounts of subcutaneous adipose tissue were more difficult to accurately estimate the triglyceride compositions due to signal influences from surrounding tissue boundaries. Therefore, a larger cohort of health volunteers should be studied further. Looking forward, we employed 2048 dynamics which lead to a single slice scan time of just under one minute. Reducing this number could lead to a faster scan time at the expense of SNR, especially if the current spatial resolution and flip angle train is kept. Furthermore, substantial work on Cramér-Rao lower bound optimization has shown promise for alternative optimal flip angle trains which can significantly reduce the required number of dynamics for the desired quantitative accuracy^80^. With strategic tradeoffs, expansion of this proposed MRF technique to abdominal applications within a single breath-hold should be achievable^81^.

### 4.4. Significance

Beyond the head and neck, triglyceride composition estimation has previously been investigated in the breast^82–84^, liver^85–87^, cardiac tissue^88–90^, knee^91^, thigh^92^, and bone marrow^93^. In particular, applying this methodology to the bone marrow to separate yellow from red bone marrow^94^ can potentially be applied for red bone marrow sparing to minimize dose to radiosensitive lymphocytes^95,96^, thus reducing the risk of lymphopenia^97,98^. Additionally, data from over 33,000 patients in the UK Biobank^99^ has shown significant promise for the incorporation of triglyceride composition estimation to aid in risk stratification across genders and diets^100^. Recent work has taken this further by investigating ω-3 fatty acid mapping^101^ which is critical in inflammatory pathways^102^ and may be translated to future longitudinal radiation therapy studies^103,104^. One interesting study showed that radiation therapy in myxoid liposarcomas resulted in a reduced ndb in the tumor in patients with good response^105^ indicating the possibility of effective double bond breakage from the radiation^106^. A corroborative mouse study indicated changes in SFA and PUFA in response to radiation therapy^107^.

A promising application of this theory is visualizing the MRI-visible lipids in the tumor microenvironment^108,109^ in an attempt to peer into long standing knowledge about the import role of lipids in cancer^110^. One study showed this through the substantial increase in PUFA seen following radiation therapy in a xenograft model of non-Hodgkin’s diffuse large B cell lymphoma, theorized to signal radiation therapy-induced apoptosis^111,112^. This correlation has also been seen in glioma in response to necrosis^113,114^ while also being seen in response to gene therapy^115^ and chemotherapy^116^. In addition, the PDFF values have been demonstrated as a biomarker for bone marrow cellularity^117^ which can be applied to assess bone metastases and osteoradionecrosis^118^. With the clinical applications of the MR-Linac expanding rapidly, applying the proposed MRF sequence in this study has the potential to generate novel insights into how tumors respond to radiation therapy at a high temporal density.

Simultaneous quantification of multiple MRI parameters in the same sequence is valuable on the MR-Linac where time constraints are necessary to minimize time the patient is on the table due to the combined imaging and radiation therapy delivery. Future applications of this work include: (1) expanding the proposed MRF sequence to other anatomical areas such as the prostate to longitudinally assess T1 and T2 changes^119^, (2) exploring the applications of quantitative R2* mapping for tracking hypoxia status^120^, and (3) investigating the changes in PDFF, and potentially the triglyceride composition, throughout the course of radiation therapy on the MR-Linac^121–124^. Therefore, this work should be viewed as only an introduction into what is possible on the MR-Linac and is only the surface of the future potential of its utility in optimally guiding patient care.

## Contributor Roles Taxonomy (CRediT) Attribution Statement

*Conceptualization*: L.M. and J.O.; *Data curation*: L.M. and J.O.; *Formal analysis*: L.M. and J.O.; *Funding acquisition*: C.D.F.; *Investigation*: L.M. and J.O.; *Methodology*: L.M. and J.O.; *Project administration*: J.O.; *Resources*: L.M., E.S., S.L.M., B.A.T., K.H., and J.O.; *Software*: L.M. and J.O.; *Supervision*: C.D.F. and J.O.; *Validation*: L.M. and J.O.; *Visualization*: L.M. and J.O.; *Writing – original draft*: L.M. and J.O.; *Writing - review & editing*: L.M., E.S., S.L.M., B.A.T., K.H., C.D.F., and J.O.;

## IRB Statement

All participants provided written informed consent. Volunteers were consented to an internal volunteer imaging protocol (PA15-0418), both approved by the institutional review board at The University of Texas MD Anderson Cancer Center.

## Data Availability Statement

All relevant anonymized imaging data necessary for figure reproduction are available at the following FigShare DOI: 10.6084/m9.figshare.30779273.

## Acknowledgements

None.

## Supplementary Materials

**Figure S1.**
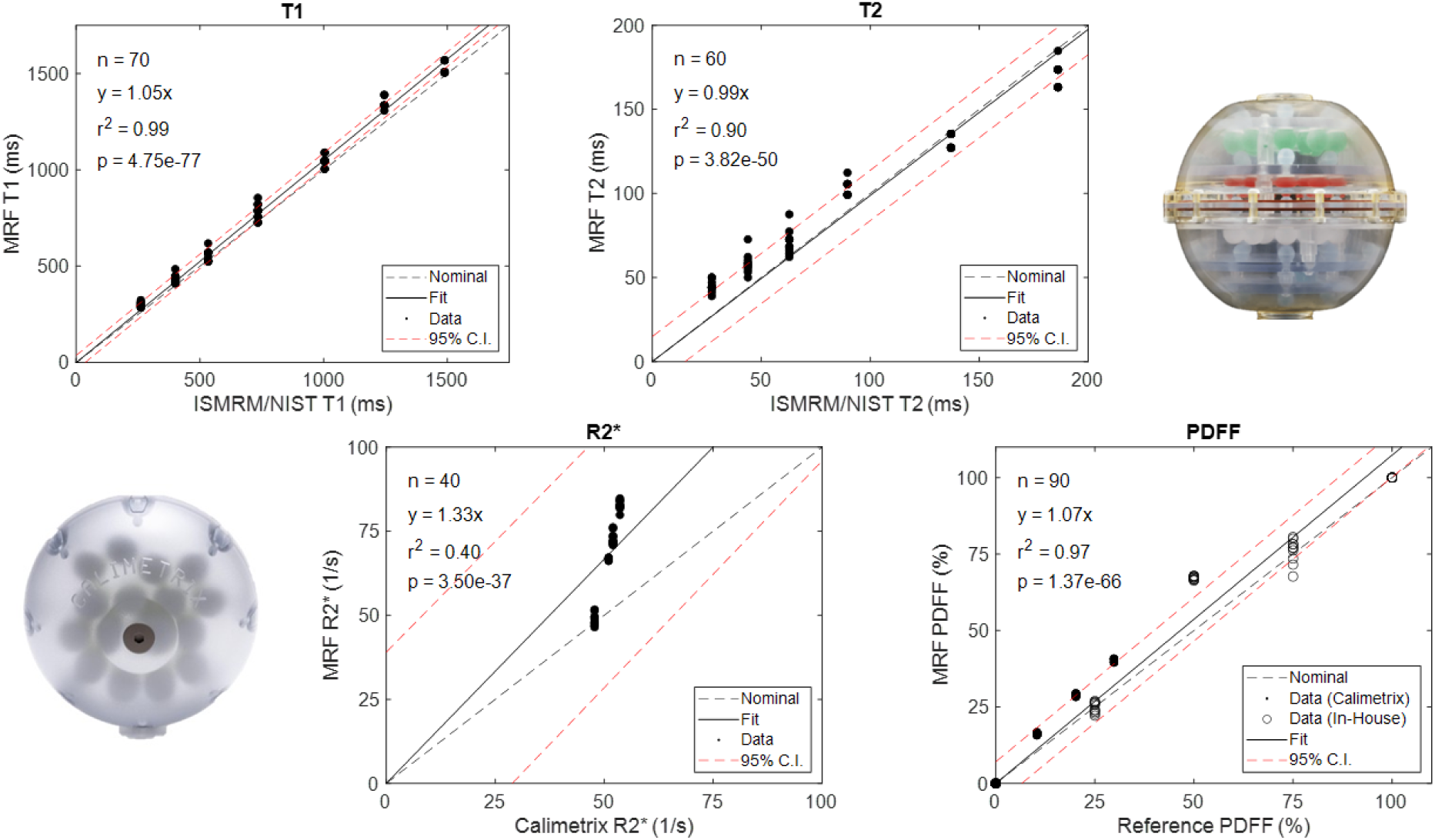
Concordance plots of the quantitative T1 (top left), T2 (top right), R2* (bottom left), and PDFF (bottom right) values from the proposed MRF sequence compared to the phantom nominal reference values (dashed black line, CaliberMRI ISMRM/NIST Model 130 for T1 and T2, Calimetrix Model 725 for R2* and PDFF, and additional in-house phantom for PDFF). For each series of data, a linear line of best fit (solid black line) was computed along with its 95% confidence intervals (dashed red lines).

**Figure S2.**
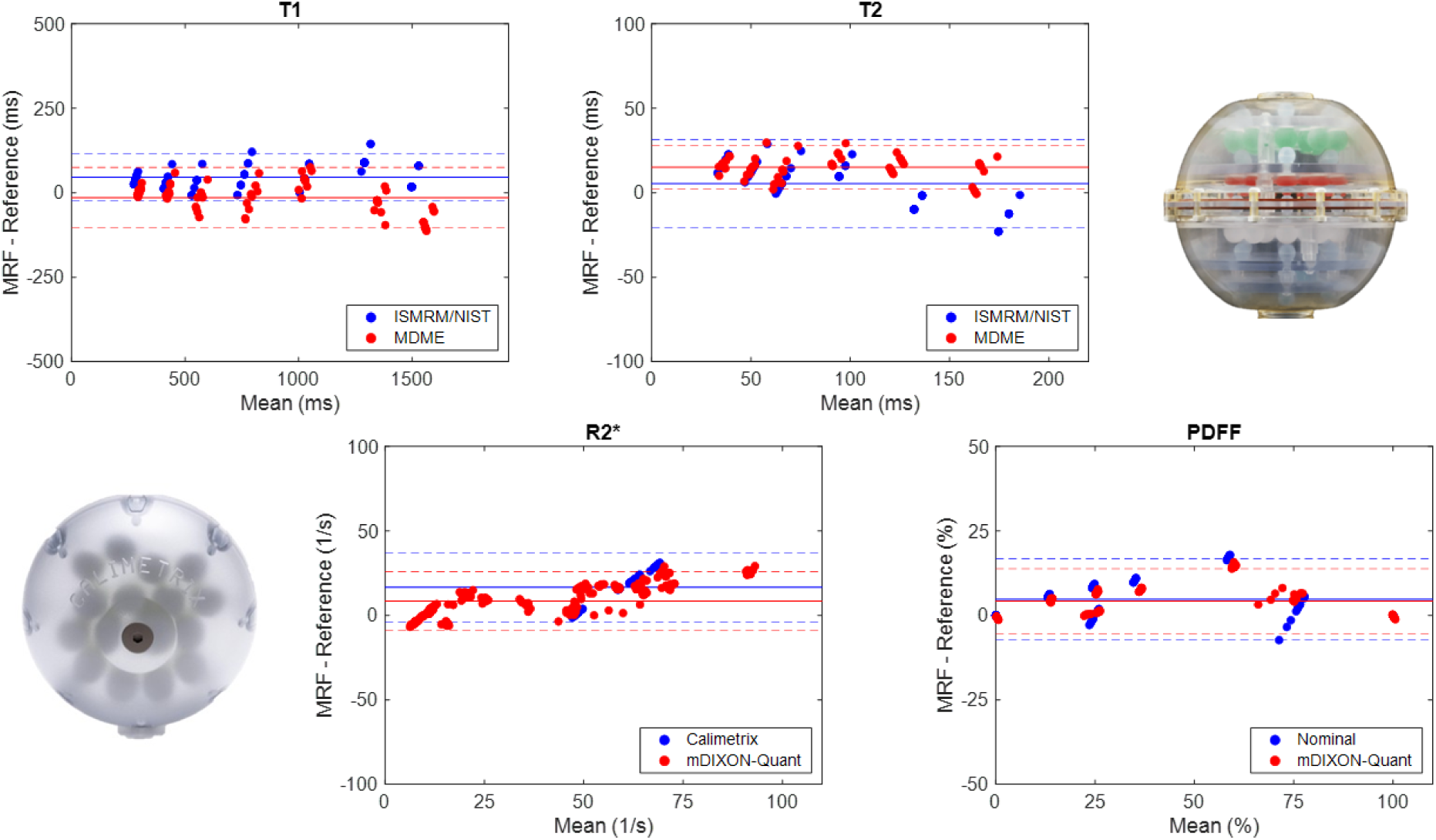
Bland-Altman plots of the quantitative T1 (top left), T2 (top right), R2* (bottom left), and PDFF (bottom right) values from the proposed MRF sequence compared to the reference values (CaliberMRI ISMRM/NIST Model 130 and MDME for T1 and T2, Calimetrix Model 725 and mDIXON-Quant for R2* and PDFF with additional in-house phantom for PDFF). For each series of data, the mean (solid line) was computed along with its 95% limits of agreement (dashed lines).

